# Development and validation of the Food Expectation Questionnaire (FEX-Q) to assess food-related perceptions and symptom expectations

**DOI:** 10.64898/2026.03.10.26348014

**Authors:** Ryo Katsumata, Ines Trindade, Stine Störsrud, Magnus Simrén, Sanna Nybacka

## Abstract

**Background:** Food-related gastrointestinal (GI) symptoms are highly prevalent in patients with IBS. Although dietary components may trigger symptoms through luminal mechanisms, cognitive expectations may also shape symptom perception within the gut-brain axis. No validated instrument currently exists to measure food-related symptom expectations. Hence, we developed and validated the Food Expectation Questionnaire (FEX-Q).

**Methods:** The FEX-Q was developed using a stepwise process including focus group interviews and face-to-face validation to ensure content validity. The finalized digital questionnaire presents 44 food images with six items rated on a visual analogue scale (VAS; 0–100), including the core item assessing food-related symptom expectation (“How severe GI symptoms do you expect after eating this food?”). Additional domains assess taste preference, willingness to eat, perceived healthiness, and perceived fat and carbohydrate content. The finalized FEX-Q was administered in a nationwide online validation survey of adults with IBS and non-IBS controls in Sweden. Participants also completed validated questionnaires including GI symptom severity (combined GSRS), psychological distress (HADS), food-related quality of life (FR-QOL), and a screening tool for food avoidance (NIAS).

**Results:** Twenty adults with IBS and non-IBS controls participated in the face-to-face validation, resulting in a final version of the FEX-Q comprising 44 food images, which were properly identified and provided a range of macronutrient distributions and trigger foods. In the nationwide online study including 134 patients with IBS and 126 non-IBS controls, the FEX-Q demonstrated strong known-groups validity (mean symptom expectation 18.4 in controls vs 50.1 in IBS), strong construct validity (perceived vs actual fat content r=0.78, p<0.001 and carbohydrate content r=0.59, p<0.001), significant convergent validity with GI symptom severity and food-related quality of life, and high internal consistency (split-half reliability Spearman–Brown corrected r=0.88).

**Conclusion:** The FEX-Q can capture individual food-related symptom expectations to distinct food images. This reliable measurement can be useful to reveal the mechanism of food-related symptom expectations and provide clinically relevant insights for personalized dietary management

## Introduction

Food-related gastrointestinal (GI) symptoms are highly prevalent among individuals with irritable bowel syndrome (IBS)^1^, one of the most common disorders of gut–brain interaction (DGBI), affecting approximately 5% of the global population^2^. Many patients report symptom provocation after consuming specific foods, particularly items rich in fermentable carbohydrates such as legumes, onions, wheat products, or certain fruits^1^. For this reason, dietary modifications remain a central therapeutic strategy. The low fermentable oligo-, di-, monosaccharide and polyol (FODMAP) diet, the most evidence-based dietary treatment for IBS^3^, likely reduces symptoms by lowering the intestinal load of fermentable substrates, thereby reducing gas production and osmotic effects^4^. However, mechanistic insights linking food exposure to symptoms are incomplete, and reduced fermentation alone cannot fully explain the variability in symptom responses.

IBS is characterized by dysregulation of the bidirectional gut–brain axis, where cognitive and affective processes shape how visceral signals are interpreted. Psychological factors, including anxiety, stress, and prior negative food experiences, can amplify symptom perception even in the absence of physiological provocation^1, 5^. Despite this, few studies have systematically examined how individuals with IBS perceive foods, how severe symptoms they expect from different foods, or how these expectations relate to actual symptom experiences. Emerging neuroimaging data suggest that simply seeing food images can activate brain regions implicated in threat processing and interoception among individuals with functional GI disorders^6^, highlighting the importance of understanding visual, cognitive, and emotional responses to food.

So far, no validated questionnaire currently exists to evaluate food-related symptom expectations, perceived nutritional characteristics, and emotional or sensory food responses in IBS. To address this gap, we developed the Food Expectation Questionnaire (FEX-Q), a standardized tool using food images to assess taste preference, emotional responses, perceived healthiness, macro-nutrient estimations, and expected severity of GI symptoms after consuming each food.

This paper describes the development process and validation results of the FEX-Q.

## Methods

### Study overview

The FEX-Q was developed and validated as part of the project *Good Gut Feelings – Food Expectations in the Brain–Gut Axis*, conducted at Sahlgrenska University Hospital, Gothenburg, Sweden. The aim of this part was to create an image-based digital questionnaire capable of reliably assessing food-related perceptions and symptom expectations in individuals with and without IBS. The finalized questionnaire is intended for use in several ongoing and future studies, including large-scale cross-sectional investigations and neuroimaging projects.

The study was approved by the Swedish Ethical Review Authority (Dnr 2025-02417-01). All participants received written information about the study and provided informed consent prior to participation.

### Original questionnaire concept

The FEX-Q presents different food images and asks participants to rate the following domains for each image:

1) taste preference
2) expected gastrointestinal (GI) symptom severity
3) emotional response to the image
4) perceived healthiness
5) perceived fat content
6) perceived carbohydrate content

All items are rated on a visual analogue scale (VAS) ranging from 0 to 100. A parallel version for healthcare professionals included items 4, 5 and 6, and replaced the symptom expectation item (2) to address whether they believe that patients with IBS would have symptoms after consumption of the foods displayed.

### Development of the initial item pool

#### Selection of food images

An initial set of 48 food images was assembled based on previous literature and clinical experience by a multidisciplinary team consisting of dietitians (SN and SS) and gastroenterologists (RK and MS). The images were selected to represent a broad spectrum of foods, including common “trigger foods” (e.g., onion-rich dishes, legumes, coffee), perceived “safe foods” (e.g., rice, berries, bananas), foods varying in fat, carbohydrate, fiber, and FODMAP content, and culturally relevant foods commonly consumed in Sweden.

#### Focus group interviews

To ensure patient relevance and content validity, a focus group consisting of ten individuals with IBS (n = 10) from our outpatient clinic was recruited. Participants were presented with the initial image set and were asked to identify the food items on the pictures, and whether they considered any additional foods to be typical symptom-triggering or symptom-safe to be included in the set. Based on their feedback, the image pool was revised and as a result, eight food images were replaced, and three additional food images were added to the pool, resulting in a total of 51 images for the preliminary questionnaire.

### Face-to-Face Validation Study

The primary aim of the face-to-face validation was to ensure that the FEX-Q is understandable, that the questions were interpreted as intended, and that the food images were perceived as relevant by participants. Participant feedback obtained during the face-to-face validation was used to revise and improve the food images and questionnaire items.

### Participants – face-to-face validation

Twenty adult volunteers participated in the face-to-face validation, including six individuals with IBS (defined according to the Rome IV diagnostic criteria) and fourteen healthy controls. This sample size was considered sufficient for qualitative validation, as data saturation in qualitative interviewing studies is typically achieved with 12–20 participants.

### Procedures – face-to-face validation

Participants completed the draft FEX-Q using a laptop or desktop monitor in the presence of a trained researcher. During completion, the researcher conducted cognitive probing to assess participants’ understanding of each component. The face-to-face validation focused on three key dimensions:

1) Food recognition: “Which food item or dish do you identify in this image?”
2) Interpretation of the questions: “How do you interpret this question?” and “What does ‘GI symptoms’ mean to you in this context?”
3) Symptom rationale: “What in this food image do you expect could provoke symptoms?”

### Evaluation Criteria – face-to-face validation

Each item was evaluated with regard to clarity of the food image, correct identification of the food, comprehensibility of the question, logical relationship between the food and perceived symptom triggers, and redundancy or overlap between items.

Responses were documented verbatim. Items that were repeatedly misunderstood, difficult to identify, or consistently perceived as ambiguous were flagged and those with recognition rates below 70% during the validation process were replaced with clearer alternatives or removed from the set.

To allow assessment of response consistency and attentiveness, one duplicate image pair was intentionally included in the questionnaire. The duplicate items were presented at different positions within the questionnaire.

### Online survey

Following finalization of the questionnaire, an online survey was conducted using the FEX-Q together with existing validated questionnaires. Study data were collected and managed using REDCap (version 16.0.13) electronic data capture tools hosted at University of Gothenburg^7, 8^. REDCap (Research Electronic Data Capture) is a secure, web-based software platform designed to support data capture for research studies, providing 1) an intuitive interface for validated data capture; 2) audit trails for tracking data manipulation and export procedures; 3) automated export procedures for seamless data downloads to common statistical packages; and 4) procedures for data integration and interoperability with external sources.

Participants were instructed to complete the survey using a laptop or desktop computer to ensure adequate visibility of the food images. They were also encouraged to use the “save and return” function if they experienced fatigue or reduced attention during survey completion.

### Participants – online survey

Adults with physician-diagnosed irritable bowel syndrome (IBS) and non-IBS controls were recruited by advertisement in social media platforms between October 17, 2025 and November 4, 2025. All participants were required to be able to understand Swedish in order to complete the survey. Non-IBS controls were defined as adults without any diseases or disorders listed in the exclusion criteria. Participants were recruited nationwide across Sweden, enhancing the heterogeneity and generalizability of the validation sample.

Exclusion criteria for IBS patients included: a history of other gastrointestinal diseases; severe psychiatric disorders, including body image–driven eating disorders or substance abuse; self-diagnosed IBS; severe active cardiac, pulmonary, neurological, or systemic inflammatory diseases (e.g., rheumatoid arthritis); and pregnancy or breastfeeding.

Exclusion criteria for non-IBS controls included: any history of gastrointestinal disease; severe psychiatric disorders, including body image–driven eating disorders or substance abuse; severe active cardiac, pulmonary, neurological, or systemic inflammatory diseases (e.g., rheumatoid arthritis); and pregnancy or breastfeeding.

Participants were provided with two movie tickets as compensation for completing the survey.

### Sample size – online survey

As this was a questionnaire development and validation study, no formal a priori sample size calculation was performed. A target sample size of approximately 100 participants per group was considered sufficient to evaluate known-groups validity, convergent validity, and reliability, and is comparable to sample sizes used in previous validation studies of patient-reported outcome measures.

### Additional questionnaires

In addition to the FEX-Q, participants completed a set of validated questionnaires to assess demographic characteristics, gastrointestinal symptoms, psychological distress, and eating-related behaviors:

#### Demographics and dietary habits

Participants provided information on socio-demographic variables, including age, sex, height, weight, education level, and occupation. Additional questions assessed general dietary habits and patterns relevant to food intake and symptom experiences.

Gastrointestinal Symptom Rating Scale – GSRS^9^ and GSRS IBS version (GSRS-IBS)^10^ The GSRS and GSRS-IBS questionnaires were used to evaluate the severity of gastrointestinal symptoms during the past week. In this study, the GSRS and GSRS-IBS questionnaires were combined into a single instrument (combined GSRS). These instruments assess a range of GI symptoms, including abdominal pain, bloating, diarrhea, constipation, and reflux-related symptoms.

#### Irritable Bowel Syndrome Severity Scoring System (IBS-SSS)^11^

The IBS-SSS is a validated instrument used to assess IBS symptom severity over the past 10 days. It includes five domains (pain severity, pain frequency, bloating, bowel habit dissatisfaction, and life interference), yielding a total score ranging from 0 to 500.

#### Hospital Anxiety and Depression Scale (HADS)^12^

The HADS is a 14-item questionnaire designed to assess psychological distress in medical populations. It provides separate subscale scores for anxiety (HADS-A) and depression (HADS-D), which were calculated and used in the analyses.

#### Patient Health Questionnaire-15 (PHQ-15)^13^

The PHQ-15 is a 15-item measure assessing the extent to which individuals are bothered by common somatic symptoms. It includes both gastrointestinal and non-gastrointestinal symptoms and provides an overall index of somatic symptom burden.

#### Food-related Quality of Life (FR-QOL-29)^14^

The FR-QOL-29 is a 29-item questionnaire that evaluates quality of life specifically related to food and eating. It assesses the impact of gastrointestinal symptoms on daily eating behaviors and food-related well-being.

#### Nine-Item Avoidant/Restrictive Food Intake Disorder Screen (NIAS)^15^

The NIAS is a 9-item screening tool for avoidant/restrictive food intake disorder (ARFID). It consists of three subscales assessing picky eating, low appetite, and fear of aversive consequences related to eating.

### Quality check – online survey

Participants with substantial missing data (>10% of items) were excluded from the analysis. Data quality was further assessed by examining internal consistency between conceptually similar items across questionnaires. Specifically, abdominal pain–related items from the combined GSRS, IBS-SSS, and PHQ were compared. Responses showing marked discrepancies between these measures were considered unreliable. For this purpose, responses indicating “no symptom” were defined as a score of 1 on the combined GSRS, 1 on the PHQ pain item, and <10 on the IBS-SSS. Conversely, responses indicating “severe symptom” were defined as scores of 6 or 7 on the combined GSRS, 3 on the PHQ, and >90 on the IBS-SSS. Participants who reported “no symptom” on one instrument but “severe symptom” on another were excluded due to inconsistency. Response times were also reviewed to identify questionnaires completed in an unreasonably short time. Surveys with an average completion time of less than 5 seconds per question were considered implausibly fast and were excluded. In addition, visual inspection of response distributions was performed, and data characterized by extreme response bias or repeated identical values across items were excluded. Finally, within-participant response variability was examined using the standard deviation across FEX-Q items. Extremely low variability suggestive of non-engaged responding was considered as an additional indicator of poor data quality.

### Outcomes – online survey

The primary outcome of the online survey was the mean food-related symptom expectation score, calculated for each participant as the average symptom expectation rating across all 44 food images.

Secondary outcomes included the composite scores for the other FEX-Q domains taste preference, perceived fat content, perceived carbohydrate content, and perceived healthiness, were analyzed as secondary outcomes. All outcome variables were analyzed both at the participant level and at the food image level, as appropriate.

### Final construct validation

#### Known-groups validity: comparison between IBS and control groups

Known-groups validity was evaluated by comparing responses between patients with IBS and non-IBS controls. Between-group differences in mean FEX-Q scores were examined for each questionnaire domain.

#### Construct validity: correspondence between perceived and actual nutrient content

To evaluate construct validity, perceived fat and carbohydrate content ratings were compared with objectively calculated nutrient content for each food image. Actual fat and carbohydrate amount per portion were obtained by the nutrient calculation software Dietist XP (kostdata.se, Stockholm, Sweden) and standardized recipes. These values were correlated with the mean perceived fat and carbohydrate ratings across participants using image-level Spearman correlation analyses.

#### Convergent validity: associations with established clinical measures

Convergent validity was assessed by examining associations between the mean food-related symptom expectation score and established validated questionnaires. Specifically, correlations were calculated between FEX-Q scores and measures of gastrointestinal symptom severity (combined GSRS), food-related quality of life (FR-QoL), and eating-related anxiety (NIAS).

#### Reliability (internal consistency)

The internal consistency of the mean food-related symptom expectation score was assessed using split-half reliability^16^. Split-half reliability was evaluated using 1,000 random splits of the 44 items. In addition to simple random splits to make two halves, a stratified split-half approach was performed in which items were first categorized into high (n = 14), middle (n = 16), and low (n = 14) scoring strata. Each half was then constructed by randomly sampling an equal number of items from each stratum.

## Statistical analysis

Continuous variables are presented as mean ± standard deviation (SD) or as median with interquartile range (IQR), as appropriate. Between-group comparisons between patients with IBS and non-IBS controls were performed using the Mann–Whitney U test. Associations between continuous variables were assessed using Spearman’s rank correlation coefficient. Split-half reliability was adjusted using the Spearman–Brown prophecy formula.

All statistical tests were two-tailed, and a p-value <0.05 was considered statistically significant. All analyses were conducted using SPSS Statistics version 29.0.1.1 (IBM Corp., Armonk, NY, USA) and MATLAB R2022b (MathWorks Inc., Natick, MA, USA).

## Results

### Face-to-face validation phase

#### Participant Characteristics

Twenty adults participated in the face-to-face validation, with mean age of 46.2 years± 16.3, and 65% females. Participant characteristics are summarized in Supplementary Table S1.

#### Food recognition

All 44 food images included in the final version of the FEX-Q were correctly identified by all participants.

#### Question Comprehension

All participants found the VAS questions intuitive and easy to understand. Minor wording adjustments were made to improve clarity and consistency. For example, some participants expressed uncertainty about the term “gastrointestinal symptoms.” In response, an explanatory statement was added to the first page of the questionnaire, clarifying that GI symptoms include events such as abdominal pain, gas, bloating, and heartburn symptoms.

Regarding the item assessing emotional response to the food images, several participants reported that their answers were primarily driven by taste preference rather than by an independent emotional reaction to the image. This interpretation was supported by a significant positive correlation between the emotional response item and taste preference ratings (r = 0.37, p = 0.007), indicating conceptual overlap between the two constructs. Based on participants’ feedback that willingness to eat varied considerably across foods and was conceptually distinct from preference, a new item was introduced: “How likely are you to eat this food?”.

#### Response consistency

The correlation between the two presentations of the duplicate item was high (e.g., r = 0.77, p<0.001).

#### Symptom Expectation Reasoning

Participants based their symptom expectations on recognizable food characteristics, most commonly on FODMAP content, fat content, portion size, and prior individual experiences. This supports the content validity of the FEX-Q by demonstrating that responses were grounded in meaningful and clinically relevant reasoning.

#### Final FEX-Q Structure

The final version of the FEX-Q that was used in the online survey consisted of 44 food images, each rated using six visual analogue scale (VAS) items:

Q1. Taste preference (“How do you like the taste of this food?”)

Q2. Expected gastrointestinal (GI) symptom severity (“How severe gastrointestinal symptoms do you expect after eating this food?”)

Q3. Willingness to eat (“How likely are you to eat this food?”)

Q4. Perceived healthiness (“How healthy do you perceive this food?”)

Q5. Perceived fat content (“How much fat do you think this food contain?”)

Q6. Perceived carbohydrate content (“How much carbohydrates do you think this food contain?”)

Optional items:

Q7. Emotional response (“Which emotions do this food evoke?”)

Q8. Healthcare workers perceptions (“How much gastrointestinal symptoms do you believe patients with IBS will have after eating this food?”)

An item assessing emotional response to the food images was included in earlier development stages but was not included in the final online version due to conceptual overlap with taste preference identified during the face-to-face validation.

### Online survey

#### Participant Characteristics

A total of 332 individuals provided informed consent and initiated the online survey. Of these, 42 participants did not complete the questionnaire and were thus excluded. Based on response time, 28 were excluded due to the short response time. In addition, two participants were excluded during data quality assessment due to marked inconsistencies between conceptually similar abdominal pain items. In total, 260 participants were included in the final analysis.

This comprised 134 patients with IBS (median age 41 years, 95% female) and 126 non-IBS controls (median age 38 years, 79% female). Demographic and clinical characteristics of each group are presented in Table 1.

**Table 1.** Demographic and clinical characteristics of participants from the online survey.

|  | **Non-IBS control**<br>n=126 | **IBS**<br>n=134 | ***p-value*** |
| --- | --- | --- | --- |
| **Age (years), median [IQR]** | 38 [28-53] | 41 [31-53] | 0.23 |
| **Female, n (%)** | 99 (78.6%) | 127 (94.8%) | <0.001 |
| **BMI (kg/m<sup>2</sup>), median [IQR]** | 23.3 [21.6-26.2] | 24.0 [21.3-27.6] | 0.74 |
| **IBS subtype, n (%)** |  |  |  |
| IBS-C | - | 38 (28.4) | - |
| IBS-D | - | 38 (28.4) | - |
| IBS-M | - | 38 (28.4) | - |
| IBS-U | - | 20 (14.9) | - |
| **IBS-SSS median [IQR]** |  | 305 [224.5-352.5] |  |

IBS, irritable bowel syndrome; IQR, interquartile range; BMI, body mass index; IBS-C, constipation predominant IBS; IBS-D, diarrhea predominant IBS; IBS-M, mixed type IBS; IBS-U, unsubtyped IBS; IBS-SSS, irritable bowel severity scoring system.

#### Known-groups validity

Between-group comparisons revealed significant differences between patients with IBS and controls for Q1–Q4. The largest difference was observed for symptom expectation (Q2), with patients with IBS reporting substantially higher expected symptom severity than controls (controls: 18.4 ± 15.3 vs IBS: 50.1 ± 12.3). Significant differences were also found for willingness to eat (Q3) (controls: 67.2 ± 11.5 vs IBS: 55.5 ± 11.9). Smaller, but still statistically significant differences were observed for taste preference (Q1) (controls: 69.0 ± 9.6 vs IBS: 64.8 ± 10.1) and perceived healthiness (Q4) (controls: 58.3 ± 7.4 vs IBS: 56.3 ± 6.6). In contrast, no significant differences were detected between groups for perceived fat content (Q5) (controls: 41.1 ± 7.9 vs IBS: 43.2 ± 8.1) or perceived carbohydrate content (Q6) (controls: 57.9 ± 10.8 vs IBS: 57.3 ± 9.9). Mean values and standard deviations for all questionnaire items are provided in Supplementary Table S2.

#### Construct validity: correspondence between perceived and actual nutrient content

At the food image level, perceived fat and carbohydrate content showed strong positive correlations with objectively calculated nutrient amounts per portion in both patients with IBS and controls (Figure 1). Perceived fat content was strongly correlated with actual fat content in both groups (IBS: r = 0.78, p<0.001; controls: r = 0.78, p<0.001; Figure 1A). Similarly, perceived carbohydrate content correlated significantly with actual carbohydrate content in both patients with IBS (r = 0.59, p<0.001) and controls (r = 0.59, p<0.001) (Figure 1B), supporting good construct validity of the FEX-Q nutrient perception items.

**Figure 1.**
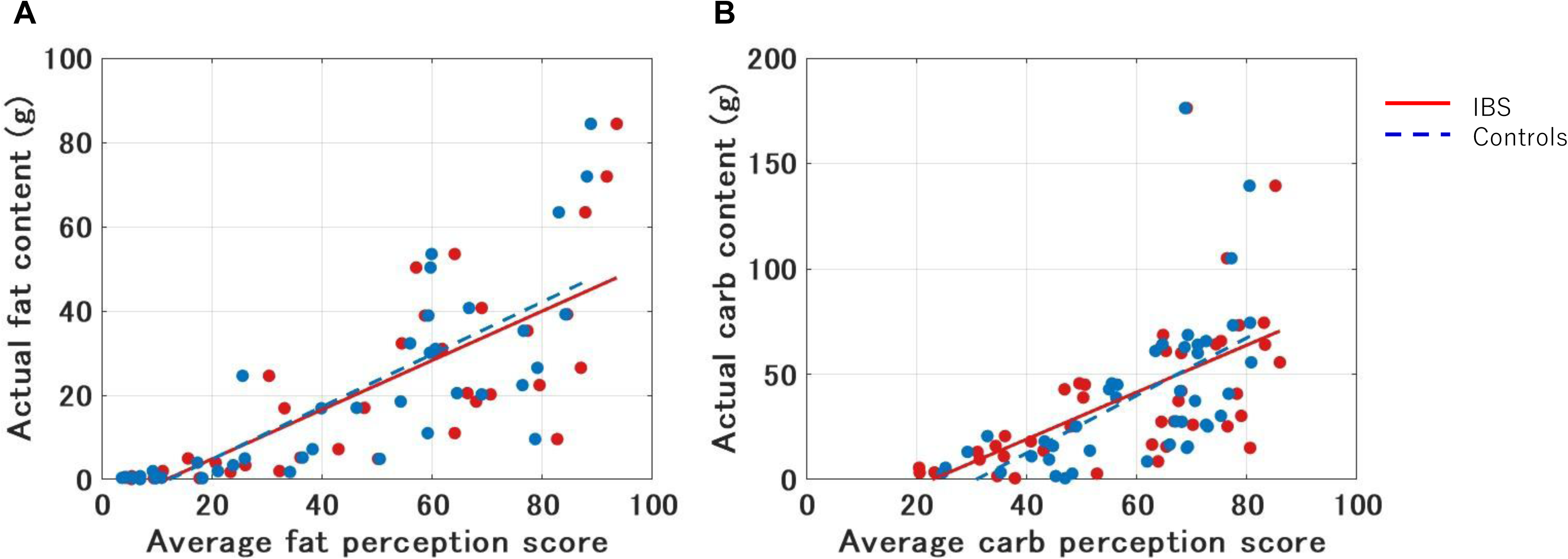
Correspondence between perceived and actual nutrient content. (A) Correlation between actual fat content and average fat perception score in both patients with IBS and non-IBS controls. (B) Correlation between actual carbohydrate content and average carbohydrate perception score in both patients with IBS and non-IBS controls.

#### Convergent validity: associations with other variables

The mean food-related symptom expectation score showed significant positive correlations with gastrointestinal symptom severity (combined GSRS; r = 0.42, p <0.001) and impaired food-related quality of life (FR-QoL; r =-0.39, p <0.001) among patients with IBS. Moderate associations were also observed with measures of psychological distress, including HADS and PHQ scores, as well as with eating-related anxiety assessed by the NIAS. The associations are illustrated in Figure 2.

**Figure 2.**
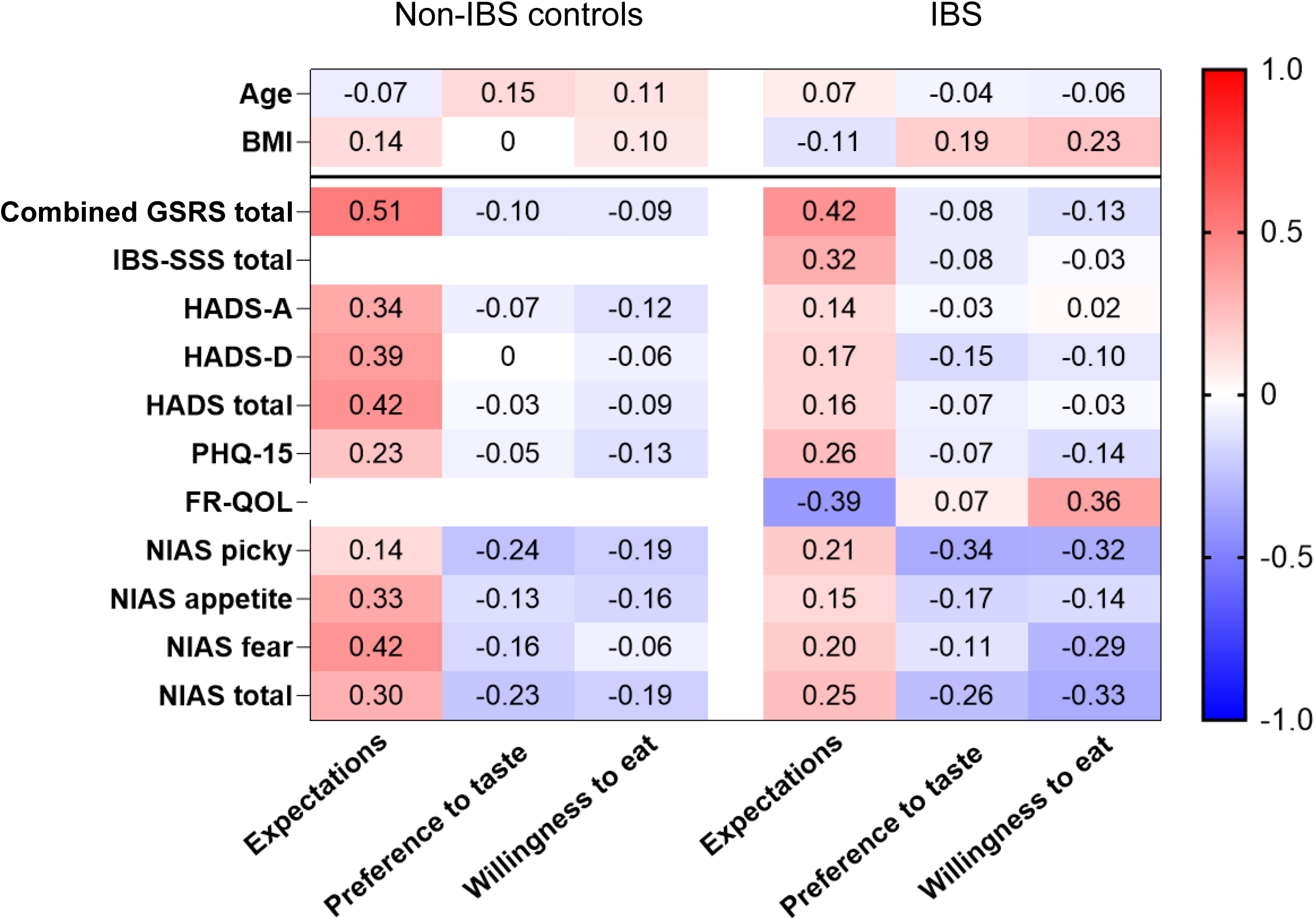
Associations between average symptom expectation scores and other clinical variables in patients with IBS and non-IBS controls. Spearman’s rank correlation coefficients (r) are visualized, with colors ranging from blue (r =-1.0) to red (r = 1.0).

#### Reliability: Internal consistency

Split-half reliability was calculated separately for IBS patients and non-IBS controls. Across 1,000 random splits, the median correlation between halves was r = 0.792 (95%CI: 0.666-0.861) in IBS patients and r = 0.785 (0.666–0.858) in controls. After Spearman–Brown correction (SB), reliability remained high in both groups (IBS: SB = 0.884; controls: SB = 0.880).

## Discussion

In this study, we developed and validated the FEX-Q, a novel image-based questionnaire designed to assess food-related perceptions and symptom expectations in individuals with IBS and in the general population. Face-to-face validation confirmed that participants could accurately recognize the presented foods, understand the questionnaire items as intended, and base their symptom expectations on identifiable food characteristics, supporting strong content validity.

The present findings provide initial evidence for the validity and reliability of the FEX-Q. Known-groups validity was demonstrated by consistently higher symptom expectation scores in patients with IBS compared with controls, in line with clinical observations that individuals with IBS frequently anticipate adverse gastrointestinal responses to food. Construct validity was supported by strong correspondence between perceived and objectively derived nutritional characteristics of food images, indicating that responses reflected meaningful interpretations rather than arbitrary judgments. Convergent validity was further supported by significant associations between symptom expectation scores and established measures of gastrointestinal symptom severity and food-related quality of life. In addition, internal consistency was confirmed by robust split-half reliability across repeated random item partitions.

The FEX-Q therefore provides a user-friendly tool to investigate food-related symptom expectations as a measurable construct. In forthcoming studies, it will be used to (1) compare expectation patterns between IBS patients and non-IBS individuals, (2) examine psychological, dietary, and clinical correlates of these expectations, and (3) evaluate how accurately healthcare professionals anticipate patients’ symptom experiences.

This study has several strengths. The FEX-Q was developed through a rigorous, stepwise process grounded in direct patient input, ensuring strong content relevance. Qualitative face-to-face interviews with participants confirmed clarity, interpretability, and ecological validity of the questionnaire items. A further strength is the relatively large validation sample, allowing robust quantitative evaluation of reliability and construct validity. The development process involved a multidisciplinary team, integrating expertise in gastroenterology, nutrition science, and behavioral research. Importantly, the instrument captures not only nutritional judgments but also visual, emotional, and cognitive dimensions of food perception, providing a more comprehensive assessment than traditional diet-based approaches.

Several limitations should be considered in this study. Firstly, the questionnaire cannot encompass all possible foods or symptom-relevant items. Instead, representative items were selected to capture core perceptual domains, which may not fully reflect individual-specific trigger foods. Further, the cross-sectional design does not allow conclusions regarding causality between symptom expectations, dietary perceptions, and clinical outcomes.

Additionally, although appropriate for qualitative validation and initial psychometric evaluation, replication in independent cohorts will be important to further strengthen external validity.

## Conclusion

The FEX-Q is a comprehensible and clinically relevant tool for assessing food-related perceptions and GI symptom expectations. It is now ready for application in clinical and mechanistic IBS research.

Future studies will examine how different perceptual domains assessed by the FEX-Q interact with objective dietary intake and symptom expectation. In addition, a separate online survey was conducted among healthcare professionals using a healthcare-adapted version of the FEX-Q. These data will be used in future work to examine how accurately healthcare professionals anticipate patients’ food-related symptom expectations and perceptions, and to explore potential gaps between patient experiences and clinical assumptions.

## Data Availability

All data produced in the present study are available upon reasonable request to the authors

