## Supplementary table for "Development and validation of the Food Expectation Questionnaire (FEX-Q) to assess food-related perceptions and symptom expectations"

| Supplementary Table 1. Demographic of participants from the face-to-face validation. | |
| --- | --- |
|  | **Participants** n=20 |
| **Age (years), median [IQR]** | 43 [31-59] |
| **Female, n (%)** | 13 (65%) |
| **BMI (kg/m^2^), median [IQR]** | 25.1 [21.0-27.0] |
| **IBS, n (%)** | 6 (30) |

| Supplementary Table 2. Mean values and standard deviation for all questions to food images | | | |
| --- | --- | --- | --- |
| Question items median (SD) | **Non-IBS control** n=126 | **IBS**  n=134 | ***p-value*** |
| Q1. Taste preference | 69.0 (9.6) | 64.8 (10.1) | ＜0.001 |
| Q2. Symptom expectation | 18.4 (15.3) | 50.1 (12.3) | ＜0.001 |
| Q3. Willingness to eat | 67.1 (11.5) | 55.5 (11.9) | ＜0.001 |
| Q4. Perceived healthiness | 58.3 (7.4) | 56.3 (6.6) | 0.01 |
| Q5. Perceived fat content | 41.1 (7.9) | 43.2 (8.1) | 0.02 |
| Q6. Perceived carbohydrate content | 57.8 (10.8) | 57.3 (9.9) | 0.34 |
